# Mobile Primary Healthcare for post-COVID Patients in Rural Areas: a Proof-of-Concept Study

**DOI:** 10.1101/2022.04.26.22274329

**Authors:** Andreas Stallmach, Katrin Katzer, Bianca Besteher, Kathrin Finke, Benjamin Giszas, Yvonne Gremme, Rami Abou Hamdan, Katja Lehmann-Pohl, Maximilian Legen, Jan Christoph Lewejohann, Marlene Machnik, Majd Moshmosh Alsabbagh, Luisa Nardini, Christian Puta, Zoe Stallmach, Philipp A. Reuken

## Abstract

**Introduction:** Post-COVID syndrome is increasingly recognized as a new clinical entity after SARS-CoV-2 infection. Patients living in rural areas may have to travel long with subjectively great effort to be examined using all necessary interdisciplinary tools. This problem could be addressed with mobile outpatient clinics.

**Methods:** In this prospective observational study, we investigated physical fitness, fatigue, depression, cognitive dysfunction and dyspnea in patients with post-COVID syndrome in a mobile interdisciplinary post-COVID outpatient clinic. Upon referral from their primary care physician, patients were offered an appointment at a mobile post-COVID outpatient clinic close to their home.

**Results:** We studied 125 patients (female, n=79; 63.2%) in our mobile unit. All patients reported symptoms lasting for more than 12 weeks after acute infection. 88.3% and 64.1% of patients reported significant impairment in physical and mental quality of life. Patients reported a median of three symptoms. The most frequently reported symptoms were fatigue (86.4%), cognitive dysfunction (85.6%), and dyspnea (37.6%). 56.0% of patients performed at <2.5^th^ percentile at the 1 min sit-to-stand test compared to age and sex-matched healthy controls and 25 patients (20.0%) exhibited a drop in oxygen saturation. A questionnaire given to each patient regarding the mobile unit revealed a very high level of patient satisfaction.

**Conclusion:** There is an increasing need for high-quality and locally available care for patients with post-COVID syndrome. A mobile post-COVID outpatient clinic is a new concept that may be particularly suitable for use in rural regions. Patients’ satisfaction following visits in such units is very high.

## Introduction

One of the most concerning aspects of the ongoing Coronavirus disease-2019 (COVID-19) pandemic is the development of serious post-infectious clinical sequelae. This phenomenon, currently known as post-COVID-19 syndrome, develops in a significant proportion of individuals who have recovered from asymptomatic or symptomatic infection with Severe Acute Respiratory Syndrome-Coronavirus-2 (SARS-CoV-2). Because post-COVID syndrome is a novel diagnosis, its full nature, frequency, and etiology remain poorly characterized.^1^ At this time, post-COVID syndrome is broadly defined as the persistence of physical and/or neuro-psychological symptoms following recovery from a confirmed or highly probable SARS-CoV-2 infection.^2^ A recent report from the World Health Organization (WHO) that utilized Delphi methodology defined post-COVID syndrome as a set of symptoms exhibited by individuals with a history of probable or confirmed SARS-CoV-2 infection that emerge within three months of the onset of acute infection and persist for at least two months thereafter that cannot be explained by an alternative diagnosis. Most importantly, the WHO Delphi panel stressed that these symptoms must be severe enough to have an impact on everyday functioning.^3^ This approach separates patients with post-COVID syndrome from individuals with persistent symptoms but who remain able to participate in their routine work and social life.^4^

Overall, more than 50 different symptoms have been reported by patients diagnosed with post-COVID syndrome.^5,6^ The most common symptoms include, but are not limited to fatigue, shortness of breath, depression, and cognitive dysfunction.^7^ Given the large number and variety of symptoms and complaints, health care provided to these patients must follow an interdisciplinary and holistic approach. This approach is often unavailable to patients who reside in remote, rural areas.

Mobile health clinics may be capable of delivering high-quality care to populations in underserved areas. Currently, an estimated 2000 mobile clinics operate across the United States and provide healthcare to seven million at-risk individuals each year.^8,9^ The results of several studies documented that health care provided by these clinics results in cost savings and improved outcomes in both children and adults.^10,11^ Mobile laboratories might provide rapid diagnoses and thus prevent the spread of infections such as SARS-CoV-2.^12^ Based on these observations, this manuscript presents the results of a proof of concept study that assessed the practicality and patient acceptance of an interdisciplinary mobile outpatient clinic specifically designed to address post-COVID syndrome. For this goal, we used a medical bus, i.e., a “mobile post-COVID-outpatient clinic on wheels” that traveled within the Free State of Thuringia. Thuringia is the fifth smallest German federal state by population (approx. 2.2million inhabitants) and includes numerous rural areas.

## Material and methods

### Mobile care unit procedures

We prospectively enrolled 125 patients who were seeking care at the post-COVID out-patient clinic of Jena University Hospital from March 14 to April 15, 2022. All patients treated in the mobile units received a structured examination that lasted approximately 2.5 hours. Upon arrival, a nurse welcomed the patient, organized the administrative tasks, and measured vital signs including height, body weight, heart rate, blood pressure, body temperature, and peripheral arterial oxygen saturation. Blood samples were obtained to assess complete blood counts, markers of renal and liver function, markers of myocardial damage, coagulation, levels of SARS-CoV-2 anti-spike antibodies, Epstein-Barr virus antibodies, and autoantibodies associated with post-COVID syndrome. Samples were stored at 4°C until the mobile unit returned to the hospital. Physical assessments included a one-minute sit-to-stand-test^13–15^; additional details are provided below. Muscle strength in each hand was assessed by repeated tests performed with a Jamar Hydraulic Hand Dynamometer^16^. Following the completion of the physical assessments, patients moved to the second room of the unit where they underwent a structured neuropsychological evaluation. Patients were provided with a series of questionnaires (i.e., Fatigue Assessment Scale [FAS]^17^, Brief Fatigue Inventory [BFI]^18^, Patient Health Questionnaire [PHQ9]^19^, and Short Form [SF]36 Health Survey^20^) designed to assess fatigue, depression, and overall quality of life (QoL). Additionally, information pertaining to cognitive function was assessed using the Montreal Cognitive Assessment (MoCA)^21^ test that was performed by a trained psychologist. The neuropsychological component of this examination required approximately 45 minutes; additional details are provided below. The patients were then transferred to a third room where they were interviewed by a physician who collected anamnestic information focused on the course of the acute COVID-19, the precise nature of the symptoms experienced, specific comorbidities, and any previous post-COVID diagnosis or treatments received. A physical examination was then performed and referrals for additional care (e.g., cardiology, pneumology, or radiology) were provided as necessary. Each patient was provided with detailed information on potential therapeutic options; if possible, appointments for necessary treatments were scheduled at that time. After the mobile unit returned to Jena each evening, the blood samples were sent to the central lab. Reports were written and sent to both the patient and the covering general practitioner. The institutional ethics committee of Friedrich-Schiller-University Jena approved the study (2020-1978-Daten) and written informed consent was waived due to the retrospective nature of the study.

### One-Minute sit-to-stand test

Before the test, the patient was asked to sit upright on a chair that had no armrests with feet placed on the ground in hip width and a 90° bend at the knees and the arms crossed in front of the thorax. Peripheral oxygen saturation and vital signs were measured by a nurse. The patient was asked to provide a subjective assessment of his/her dyspnea using the BORG scale.^22^ The patient was asked to stand and then sit again as many times as possible within a one-minute period without support provided by the arms. The number of repetitions achieved was tabulated by the nurse. Measurements of peripheral oxygen saturation, vital signs, and subjective dyspnea were then repeated one minute after finishing the test.

Diminished physical conditioning is revealed by a reduction in oxygen saturation of one to three points or more upon completion of the one-minute sit-to-stand test. Patient-directed termination of the test before the one-minute mark because of dyspnea or muscle pain, if the patient was capable of performing 12 or fewer sit-stand-cycles, or if the peripheral oxygen saturation dropped below 90% were all indicative of reduced physical conditioning.^23–25^ Data published by Strassmann et al.^14^ were used for comparisons with healthy controls.

### Peripheral muscle strength (handgrip strenght)

Handgrip strength was measured using a valid and reliable hand dynamometer (Jamar, Nottinghamshire, UK). Three trials were carried out for the dominant and non-dominant hand and the overall maximum was used for data calculation.^26,27^ Data published by Albrecht et al. were used for comparisons with healthy controls.

### Quality of life (QoL)

QoL was assessed using the German language version of the Short Form 36 (SF36), Version 2.^20^ The findings were interpreted based on the current manual and compared to the German Norm Cohort.^28^

### Neuropsychological evaluation – assessments

Patients treated in the mobile post-COVID unit underwent a structured neuropsychological assessment, including screening for fatigue (FAS and BFI)^17,18^, depression (PHQ-9, depression module of the PHQ-D)^19^, and cognitive dysfunction via MoCA screening^21^ performed by a trained psychologist. All patients were native German speakers; thus, German versions of all questionnaires and tests were performed according to their respective manuals. The severity of fatigue was defined via the BFI questionnaire, with scores that included <1, no fatigue; 1 to <4, mild fatigue; 4 to <7, moderate fatigue; and 7 to 10, severe fatigue. Depression was defined by the following responses to the PHQ-9: ≤4 points, minimal or absent; 5 to 9 points, mild; 10 to 14 points, moderate; and ≥15 points, severe. Cognitive dysfunction was defined as <26 points using the full version of MoCA.

### Patient survey and evaluation of the mobile post-COVID outpatient clinic

Patients were asked to evaluate their experiences using a specific questionnaire (five-point Likert scale with terms including “disagree”, “rather disagree”, “neutral”, “rather agree”, and “agree”. The following items were addressed: accessibility, organizational processes, privacy, hygiene, expectations, and preference for care provided at the hospital.

### Statistical analyses

Unless stated otherwise, we summarized the patient characteristics as absolute and relative frequencies for categorical variables, and as medians and first and third quartiles (Q1, Q3) for numerical variables. Relative frequencies and 95% confidence intervals (CIs) were used to present continuous data. For explorative comparisons between two patient groups, we applied Fisher’s exact test for categorical variables and the Mann-Whitney U test for numerical variables. For time-to-event comparisons, we applied the log-rank test and provided a corresponding Kaplan-Meier curve for illustration, as necessary. We applied uni- and multivariable logistic regression modeling to explore the associations between diagnostic findings (fatigue, depression, and cognitive dysfunction) and the initial severity of the SARS-CoV-2-infection (hospitalization, hospital stay, and WHO grade). For each combination of diagnostic findings and degree of initial disease severity, we adjusted for sex and age using a multivariable model with (adjusted) odds ratios (ORs) with 95% CIs. Nominal two-sided *p*-values are presented. For the analyses, we used SPSS 26 (IBM Inc, Armonk, NY, USA) and PRISM 6 (GraphPad Inc, San Diego, CA, USA).

## Results

### Patients

One hundred and twenty-five patients (n=79 females; 63.2%) presented at our mobile post-COVID outpatient clinic during the one-month study period. The median time between a polymerase chain reaction (PCR)-confirmed diagnosis of SARS-CoV-2 infection and presentation at the out-patient clinic was 421 days (Q1–Q3, 358–467 days). The median age of this patient cohort was 51 years (Q1–Q3, 41-58 years) and their median body mass index (BMI) was 28.5 kg/m^2^ (Q1–Q3, 24.4–31.6 kg/m^2^). Of these 125 patients, 20 (16.0%) were status-post hospitalization for treatment of their initial SARS-CoV-2 infection; 10 patients were treated in the intensive care unit (ICU). Patient demographics and details regarding their acute infections are presented in **Table 1**. All patients reported at least one symptom of post-COVID syndrome; the median number of symptoms was 3 (range, 1–6). The most frequently self-reported persistent symptoms were fatigue (n=108, 86.4%), cognitive dysfunction (n=104, 83.2%), and dyspnea (n=47, 37.6%). An overview of these findings is presented in **Figure 1**.

**Table 1:** Demographic characteristics of the 125 patients that received care at the mobile post-COVID units. Data are presented as median with first/third quartiles or as absolute numbers and percentages.

|  |  |
| --- | --- |
| Sex female/male (n) | 79 (63.2%)/46 (36.8%) |
| Age (years) | 51 (41–58) |
| BMI (kg/m<sup>2</sup>) | 28.5 (24.4–31.6) |
| Days since PCR diagnosis of infection | 421 (358–467) |
| Severity of initial infection (WHO grade) |  |
| 1 | 6 |
| 2 | 93 |
| 3 | 4 |
| 4 | 2 |
| 5 | 8 |
| 6 | 7 |
| 7 | 0 |
| 8 | 3 |
| Systolic blood pressure (mmHg) | 148 (136–168) |
| Heart rate (/min) | 80 (71–91) |
| Body temperature (°C) | 36.2 (35.8–36.5) |
| SpO<sub>2</sub> (%) | 98 (97–99) |

**Figure 1:**
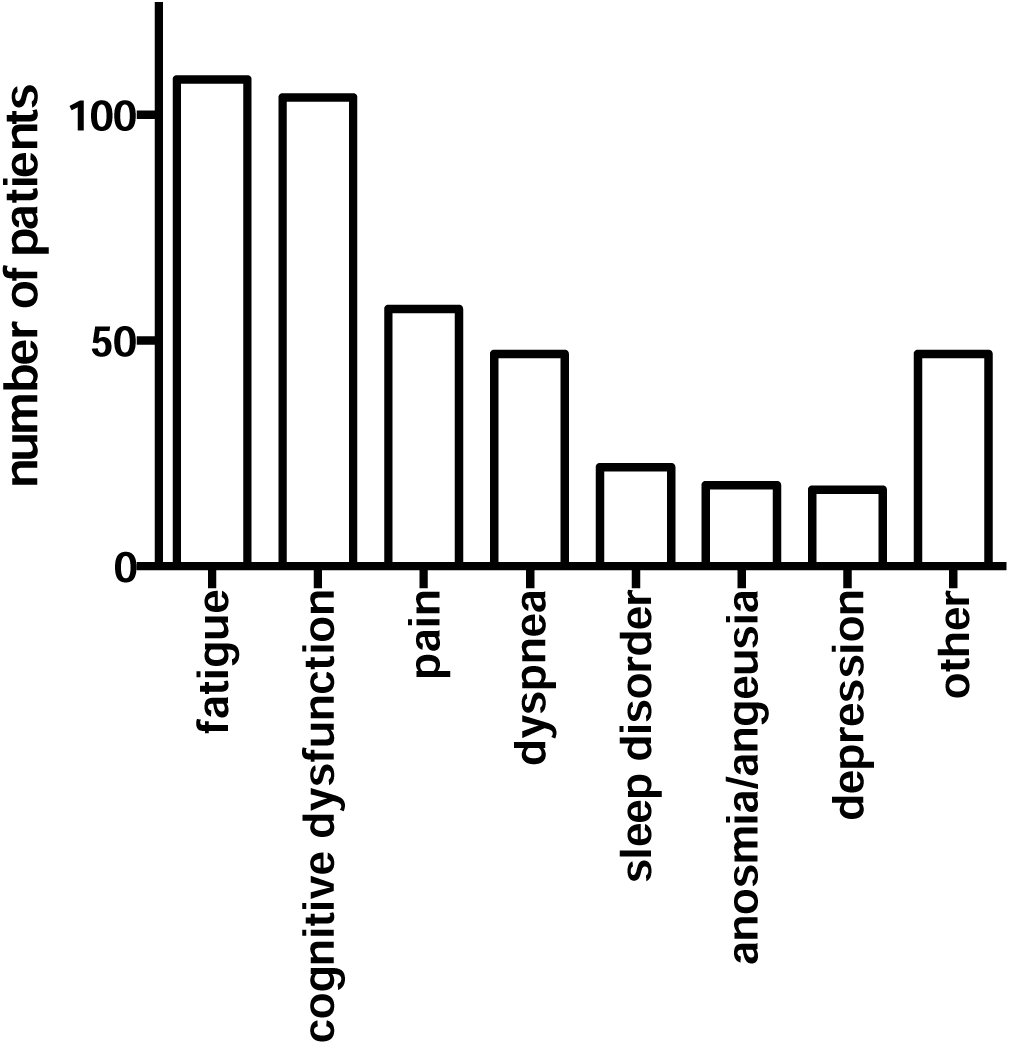
Symptoms at presentation to the mobile post-COVID healthcare unit

### Neuropsychological screening

The structured neuropsychological analysis based on patient questionnaires revealed a high prevalence of fatigue (n=117, 93.6%). Results from the BFI score revealed that 24 patients (19.2%) reported mild, 60 (48.0%) moderate, and 32 (25.6%) severe fatigue. Symptoms suggestive of depression were described by 84 patients (67.2%) in the PHQ-9, including 42 patients (33.6%) reporting symptoms indicating moderate to severe depression. As anticipated, our results revealed a broad overlap between fatigue and depression. Seventy-eight patients reported both fatigue and depression; 27 patients exclusively decribed symptoms of fatigue, while only 2 patients reported depression in the absence of fatigue (**Figure 2**). Patients with self-reported fatigue or depression were identified by structured screening approaches in 106 and 16 cases, respectively; nine additional patients with fatigue and 68 patients with depression who did not complain about fatigue or depression were subsequently identified with structured screening, while two patients reported fatigue and one reported signs of depression in the absence of specific screening results. We did not observe a correlation between pathological results in BFI, FAS and PHQ-9 with age, sex, or initial severity as indicated by WHO grade (all p>0.05).

**Figure 2:**
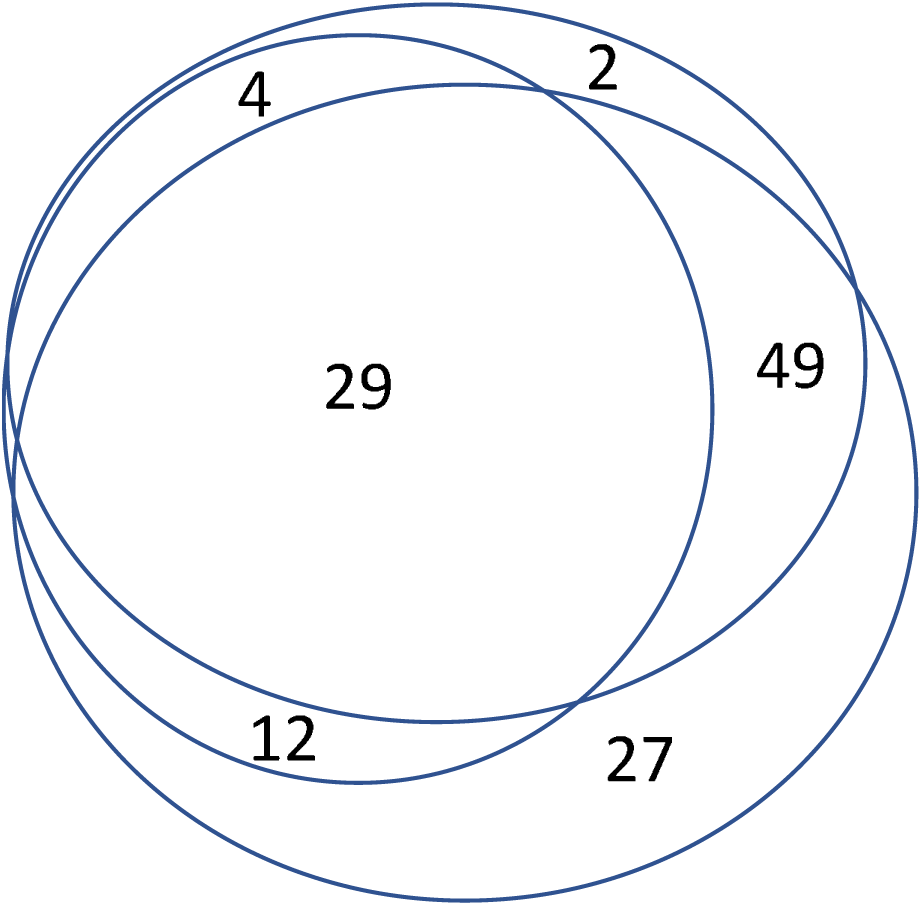
Venn diagram that indicates the overlap of patient symptoms including fatigue (blue circle), depression (green circle), and cognitive dysfunction (yellow circle).

In forty-five patients (36.0%) the MoCA test results indicated cognitive. All of these patients reported fatigue (n=12), depression (n=4), or both fatigue and depression (n=29). A pathological result in the MoCA test was not associated with sex (p=0.698) and showed a trend to be more frequent with increasing age (p=0.064) or WHO grade (p=0.08).

### Quality of life

Data addressing QoL were available for 103 of the 125 patients. Overall, the patients enrolled in our study reported impaired QoL with mean levels of 34.46 points (range, 18.01–57.62 points) in the Physical Component Summary (PCS) and 39.25 points (range, 19.26–63.96 points) in the Mental Component Summary (MCS) sum scores of the SF36. Overall, PCS and MCS sum scores <50 points were reported for 94 (91.3%) and 84 patients (81.6%), respectively. Compared to the current German Norm Cohort, significant reductions were observed in all dimensions (**Figure 3**). There were no significant differences in the reported QoL in subgroups stratified by age or sex **(Supplemental Figures 1** and **2)**.

**Figure 3:**
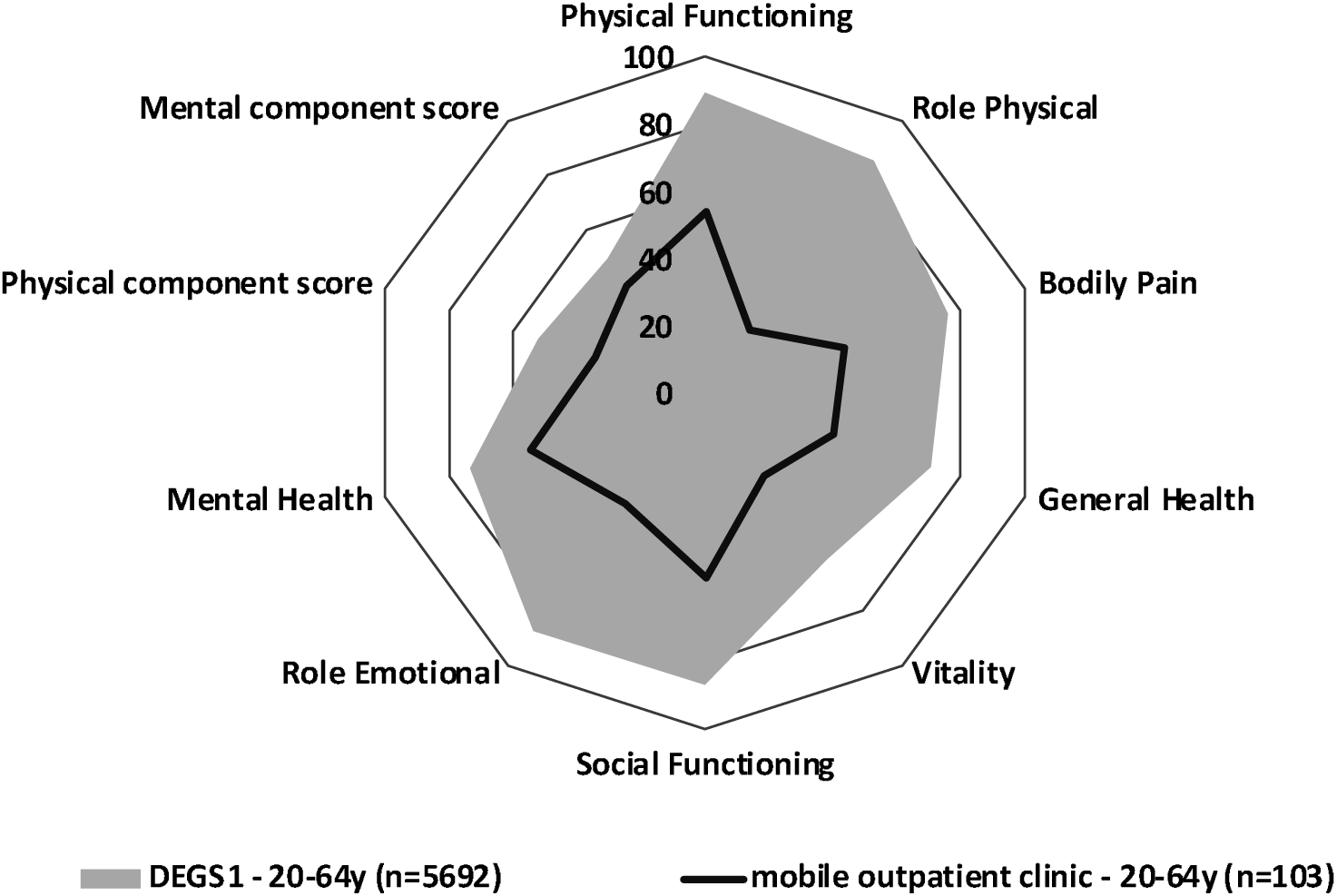
Quality of life (QoL) of patients who presented to the mobile post-COVID healthcare unit compared to the German Norm Cohort (DEGS1)

### Assessment of physical fitness

The post-COVID patients enrolled in our study performed a median number of 22 cycles (range 1–51) on the 1-minute-sit-to-stand test. Seventy of the patients (56.0%) performed at <2.5^th^ percentile compared to age and sex-matched healthy controls. A detailed distribution of the cycle count as a percentage of the age- and sex-adjusted healthy controls is shown in **Figure 4**. Lower levels of physical fitness revealed by the 1-minute-sit-to-stand tests were not associated with the initial severity of the disease (WHO grade; *p*=0.365), and no significant differences were observed when comparing hospitalized *versus* non-hospitalized patients (*p*=0.223). While poor performance on the 1-minute-sit-to-stand tests did not correlate with patient sex (*p*=1.000), we did detect a trend toward more frequent sit-to-stand cycle pathology in younger patients, although this finding did not achieve statistical significance (*p*=0.059).

**Figure 4:**
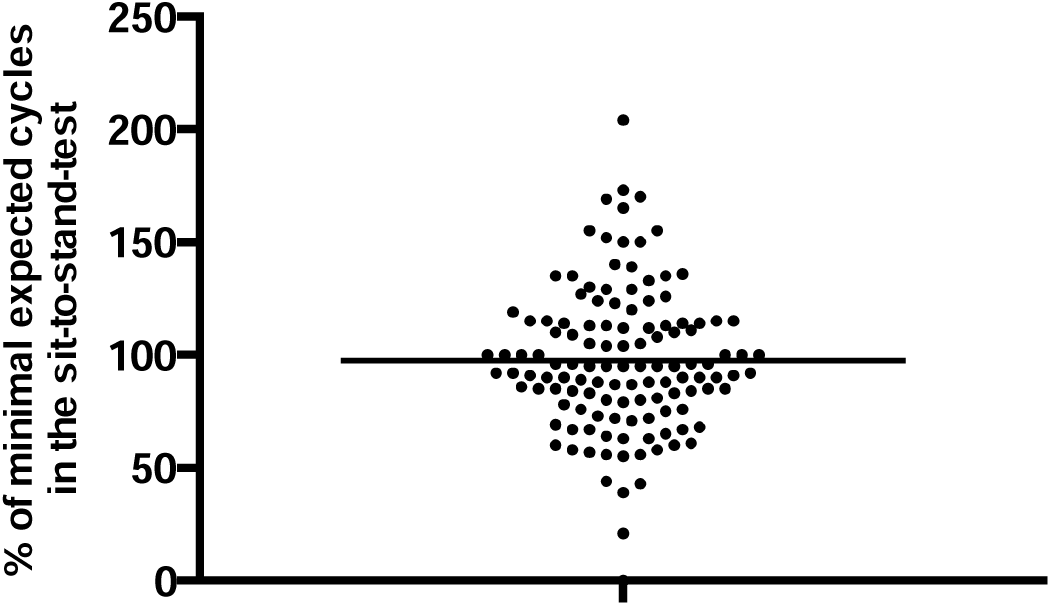
Distribution of the results of the sit-to-stand test performed on patients presenting to the mobile post-COVID healthcare unit.

Median values of self-reported dyspnea (BORG Scale) increased from 1 before the 1-minute-sit-to-stand test to 3 after the test. At the same time, the median oxygen saturation decreased from 99% (range, 93–100%) to 98% (range, 93–100%); 25 patients (20.0%) exhibited a drop in oxygen saturation of 3% or more. The patients’ heart rates increased from 82 to 105 beats per minute. Finally, median grip strength was 32.5 kg (range, 5–63kg) on the dominant side and 31.7 (range, 5.2–57.1) on the non-dominant side; these values did not differ significantly from those of the German Norm Cohort (*p*=0.203).

### Association between physical fitness, QoL, and neuropsychological symptoms

Results from the 1-minute-sit-to-stand test were not associated with QoL in the SF36 PCS sum scores (Spearman’s rho, 0.175, *p*=0.277), the MCS sum scores (Spearman’s rho, 0.119, *p*=0.199), or with any of their subdimensions. Patients reporting fatigue as per the FAS or BFI performed fewer mean sit-to-stand cycles, with the number of repetitions corresponding inversely to the increasing severity of fatigue, albeit without statistical significance (Spearman’s rho, -0.169, *p*=0.065; Spearman’s rho -0.003, *p*=0.975). Additionally, no differences in exercise-related dyspnea were reported when comparing patients with or without fatigue (*p*=0.958). Maximum grip strength was not associated with QoL as indicated by PCS and MCS Spearman’s rho, 0.072, *p*=0.472; Spearman’s rho -0.044, *p*=0.662) or with the underlying subdimensions. We also observed no differences in grip strength when comparing patients with or without fatigue (Spearman’s rho, 0.076 and 0.065, *p*=0.402 and 0.475, respectively). Additionally, no significant differences in dyspnea and oxygen saturation in patients with fatigue were observed. Additionally, symptoms suggestive of depression as indicated by PHQ-9 did not correlate with the number of sit-to-stand cycles or grip strength.

### Patient evaluations of the mobile post-COVID healthcare unit

Overall patient satisfaction with the mobile post-COVID healthcare unit was high. On a five-point-Likert scale, a mean score of 4.4 points indicated that the patients’ expectations were highly fulfilled. Additionally, the patients were satisfied with the information provided regarding the location of the unit (mean 4.6 points), reachability (mean 4.5 points), hygienic conditions within the unit (mean 4.9 points), and efforts to maintain patient privacy (mean 4.8 points). The overall organization and medical care provided were regarded as strong (mean 4 and 4.8 points, respectively). Patients reported that they had sufficient time to ask questions (mean 4.8 points). Most patients noted that they would recommend this treatment facility to friends or family members (mean 4.6 points). The only aspect associated with some ambivalence was the question that asked whether they would prefer an appointment in a mobile outpatient unit or in a more standard outpatient clinic setting (mean 2.5 points). However, when questioned directly about this issue, the most frequent response was “I take the earliest appointment I can get” (**Figure 5**).

**Figure 5:**
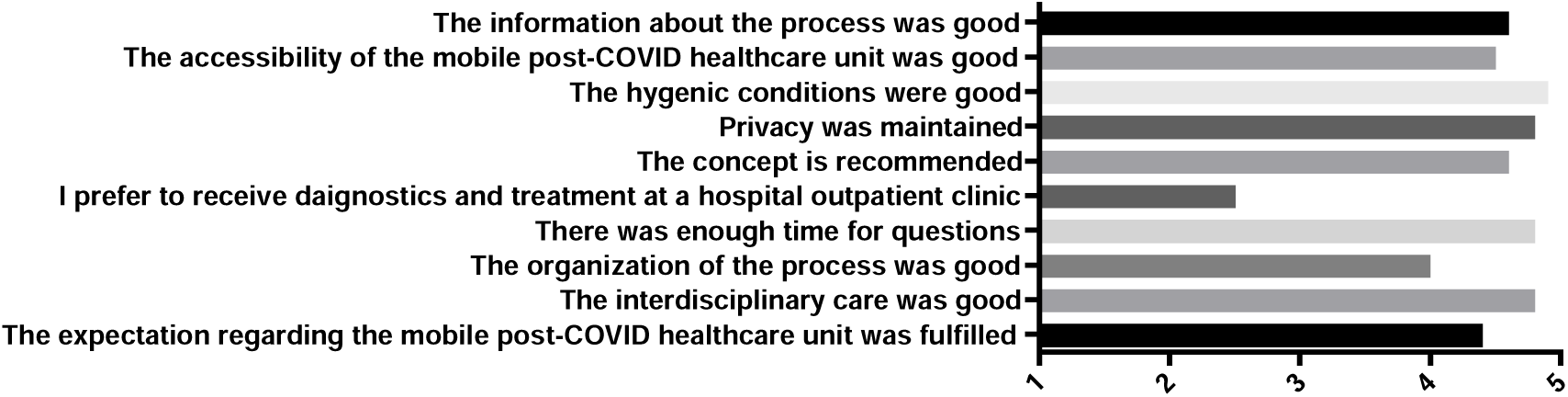
Patient evaluations of the mobile post COVID healthcare unit

## Discussion

Given the persistence and severity of the SARS-CoV-2 pandemic, Jena University Hospital opened a post-COVID outpatient clinic in August 2020. It quickly became apparent that many patients had to travel extremely long distances. Especially thos residing in rural areas often had travel times of several hours in order to obtain appropriate diagnosis and care especially.^29^ Furthermore, as the care program provided by our outpatient clinic typically required to stay in the hospital for 6–8 hours, many of the patients presenting with post-COVID syndrome became fatigued and overwhelmed. Furthermore, given the prevalence of concentration and attention disorders associated with this syndrome, many patients were simply unable to arrange a visit to the special outpatient clinic located at just one center. The goals of our mobile post-COVID health service were thus to (1) decrease barriers to obtaining interdisciplinary post-COVID care by providing an outpatient clinic experience that is close to home and easy for patients to reach, and (2) to provide all the necessary diagnostic procedures in a straightforward manner at one timepoint and within one location. Our proof-of-concept study clearly demonstrated the practicality of this approach and most important, patient acceptance of these services, most notably in rural areas of Thuringia. Mobile health clinic visits were scheduled primarily based on geography and secondarily on patients’ time preferences. Use of the mobile post-COVID healthcare units reduced the median travel distance by a median of 74 km and thus reduced the travel time by ∼1.5 hours.

The patients presenting to our mobile unit with symptoms that were similar to those reported by patients visiting our regular outpatient clinic^7^ and are thus considered “typical” post-COVID patients. The examinations performed in the mobile unit were practical given the limited resource setting and permitted us to provide the patients with detailed advice regarding efforts to improve their condition and general health. The patients appreciated the well-coordinated diagnostic processes and were overall impressed with the quantity and quality of the assessment instruments available in this setting. With respect to the cognitive evaluation, we will consider switching to a more comprehensive, digital test providing detailed subscore information on diverse cognitive functions rather than using a broad screening value provided by the overall sum score of the MoCA test. A solution could be a transition to a tablet-based test, such as the OCS-plus^30^. We believe that this option may result in further improvements in diagnostic accuracy.

Post-COVID patients frequently complain of a decrease in physical activity combined with exercise-induced shortness of breath. An assessment of these limitations makes sense when assessing prognosis and initiating therapeutic procedures. Classic imaging modalities do not assess the extent of functional limitation. While classic cardiopulmonary exercise tests are available, we chose the sit-to-stand test, which can be carried out easily and almost anywhere without specialized training or equipment. Of note, 70% of the patients in our cohort showed deviations from the age- and gender-specific norms; this finding is consistent with that reported previously by Núñez-Cortés et al.^31^ Interestingly, the degree of physical dysfunction recorded for the patients who had been hospitalized due to SARS-CoV-2 infection was not different those with mild to moderate infection that was treated on an outpatient basis. These results conflict with those reported by Núñez-Cortés et al.^31^; specifically, we observe no correlation between the results of the one-minute sit-to-stand test with initial severity of the disease. However, this may be due to the comparatively few patients in our cohort who required hospitalization with or without an ICU stay patients.

Privacy within the mobile unit was ensured by the spatial separation of the examination stations. Patient concerns were recorded in a structured manner and objective data on physical fitness, lung function, cognitive dysfunction, fatigue, and depressive symptoms were collected. The number of human resources required for this effort (i.e., doctor, psychologist, medical assistants, and mobile unit drivers) initially appear to be somewhat high. However, the same diagnostic services are typically provided in a routine outpatient by two or three specialists. This requires appointment coordination and precludes timely provision of healthcare to individuals at-risk.

The concept of a mobile outpatient clinic is not formally new. These healthcare models have been used successfully in rural regions around the world to compensate for structural deficits associated with the provision of health care to this population. In Manchester, the United Kingdom, the National Health Service (NHS) Clinical Assessment and Treatment Service offers patients access to mobile specialist healthcare professionals who can diagnose symptoms and provide recommendations for additional treatment as necessary. Similarly, a mobile outpatient clinic was used by Charité in cooperation with Deutsche Bahn to facilitate rapid and effective vaccination of refugees.^32^ The Hessen Association of Statutory Health Insurance Physicians offers a rolling family physician practice to provide primary medical care to underserved areas.^33^ A survey of local politicians from the Saxony region of Germany revealed that, according to medical assistants, mobile outpatient clinics are effective at minimizing deficits in the health care provided to all members of the population.^34^

Among the limitations of our trial, it has a nonrandomized design and lacks a control group; both of these factors may introduce the possibility of a bias. For example, many post-COVID patients experienced prolonged waits for appointments in routine outpatient clinics. Thus, the rapid availability and good accessibility of the mobile unit may have had a profound influence on the positive assessments.

In summary, our study reveals that, despite the substantial organizational challenges, the use of a mobile outpatient clinic permits us to provide care for patients with post-COVID syndrome who would otherwise have difficulties in obtaining timely interdisciplinary care. We note a very high percentage of the patients who presented to the mobile unit with significant neuropsychological and physical deficits. If care provided close to home is consistently declined, these patients might receive care via telemedicine. For example, Dalbosco-Salas et al. reported that a telemedicine-based rehabilitation program applied in primary care practice was effective at improving physical capacity, QoL, and other symptoms among COVID survivors.^35^ The use of a combination approach, including a mobile post-COVID healthcare unit and interdisciplinary telemedicine interventions might be addressed in a larger controlled clinical study.

## Data Availability

The data underlying this study are available from the corresponding author upon reasonable request

## Acknowledgments

The authors thank the authorities of the counties of Saalfeld, Sömmerda, Ilmkreis, Apolda and Altenburg, all located in the state of Thuringia, Germany, for cooperation and the possibility of offering our mobile service. Furthermore, we thank Ina Geißler for excellent organization of the patients’ appointments including the information on the exact parking place of the bus and Birgit Green, Sindy Maechler and Lisa-Marie Hahnemann for excellent nursing assistance in the mobile clinic.

## Authors contributions

AS had the original idea for the concept, AS and PAR wrote the manuscript. AS, PAR, CP, KF, JCL and BB developed the diagnostic concept. KK, KF, MMA, RAH, LN and ZS recruited the patients and performed the diagnostic procedures, ML collected the clinical data. BG and PAR performed statistical analysis.

---

**Supplemental Figure 1:**
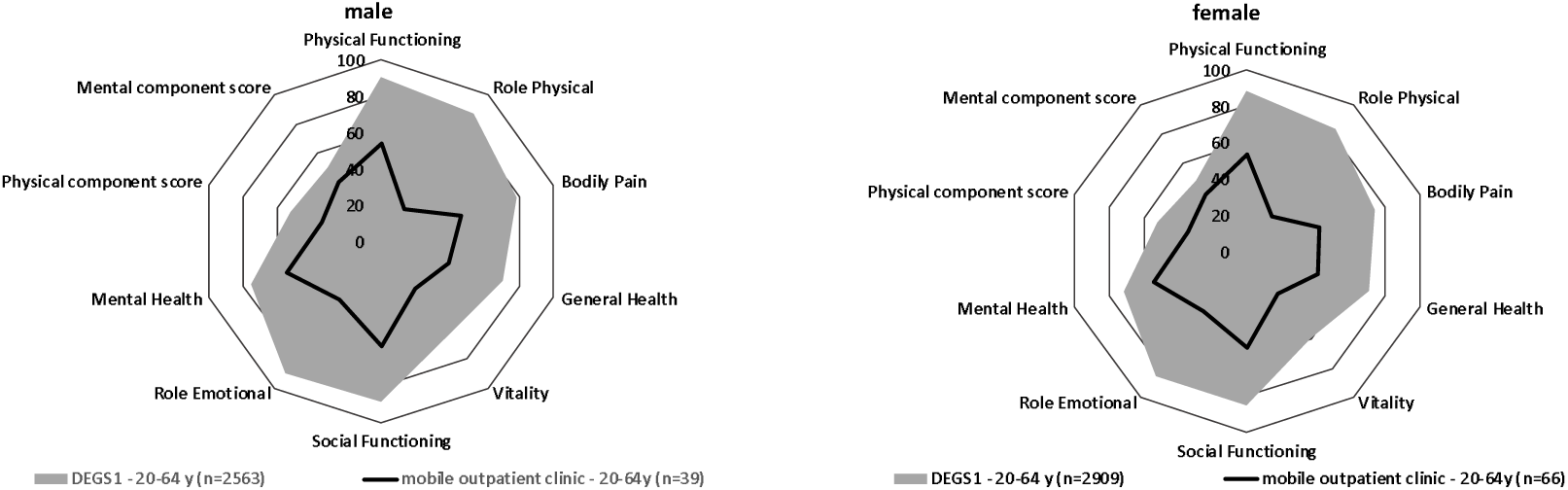
Subgroup analysis of SF36 based on patient sex

**Supplemental Figure 2:**
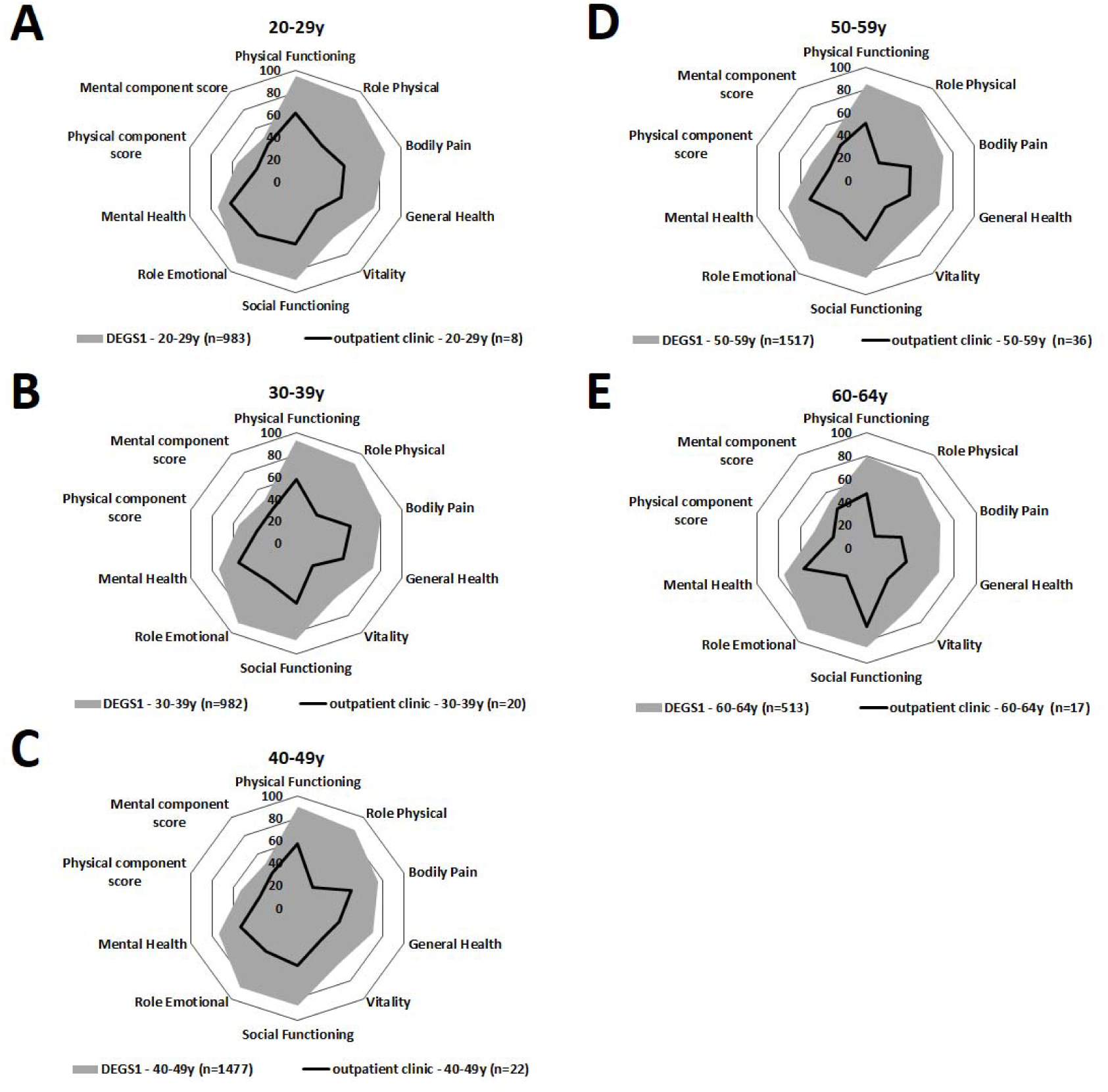
Subgroup analysis of SF36 based on patient age

